# Healthcare interventions for low-wage migrant workers: A systematic review

**DOI:** 10.1101/2024.10.26.24316170

**Authors:** Eilin Rast, Karen Lau, Rosita Chia-Yin Lin, Tharani Loganathan, Sally Hargreaves, Cathy Zimmerman, Consortium for Migrant Worker Health

## Abstract

Low-wage labour migrants often face health-damaging living and working conditions, but are frequently excluded from healthcare. The othering of migrants, bordering of healthcare and simple oversight and negligence create widening health inequalities for a society’s essential workers. This review aimed to identify the forms and effectiveness of healthcare interventions designed to make services accessible for migrant workers .

We searched for literature through Medline, Embase, Global Health, Web of Science, and Global Index Medicus (from 1 January 2000 till 9 June 2023), focussing on some of the most common sectors for forced labour (domestic work, construction, manufacturing, agriculture, mining). Primary research, reports, and grey literature from 2000 onwards containing descriptions or evaluations of healthcare interventions exclusively targeting low-wage migrant workers and their families were included. We excluded interventions focussing only on specific health conditions or disease screening. Quality appraisal was based on JBI tools. We produced a narrative synthesis separately for the interventions’ characteristics and effects. This review follows the PRISMA reporting guidelines for systematic reviews and is registered with PROSPERO (CRD42023459360).

Identified studies included 21 interventions targeting low-wage migrant workers in six countries (China, Dominican Republic, Italy, Qatar, South Africa, USA) in three sectors (agriculture, manufacturing, domestic work). Interventions included established medical facilities (e.g., general hospital care, semi-permanent primary healthcare (PHC) services); mobile clinics for PHC; and telehealth services. Interventions were provided by governmental, non-governmental, academic, and private actors. Most targeted migrant farmworkers and were primarily located in the United States. Common healthcare barriers were addressed, for example, via free care, outreach, or non-traditional hours. However, the interventions’ effects on health, access and uptake, patient satisfaction, and acceptability were largely unclear, as only six studies offered some fragmentary evaluative evidence.

Few healthcare interventions targeting migrant workers have been documented and evaluated, especially in LMICs. Although migrant workers are deemed to be mobile populations, once in the destination location, many are quite immobile when it comes to accessing healthcare. Thus, in the face of multidimensional exclusion of migrant workers, health systems cannot simply rely on the ability of this vital workforce to seek and use preventative or curative care, but healthcare services must be actively designed to be accessible to this mobile population in order to ensure health as a human right.

**Highlights:**

– **What is already known on this topic:** Many migrant workers are exposed to occupational health risks and substandard living-conditions. Due to the intersection of socioeconomic disadvantage and migrant status as well as bordering of healthcare and other services, low-wage labour migrants often face multi-dimensional exclusion from health systems. To inform policy, practice and research, we systematically reviewed evidence on targeted healthcare interventions globally for migrant workers.
– **What this study adds:** This review identified healthcare interventions for migrant workers, including: established clinics (e.g., general hospital, p healthcare centres); mobile clinics (delivering primary healthcare); and telehealth services (for chronic disease management and mental health. Common tactics to overcome exclusion from healthcare were applied via e.g., outreach, free care or language mediation. Most included interventions targeted agricultural workers.
– **How this study might affect research, practice or policy:** Findings offer several examples of approaches designed to surpass borders to healthcare commonly faced by low-wage migrant workers with prevention and treatment interventions. To improve health equity for migrant workers, budget-holders need to invest in diverse interventions that are specifically designed to reach migrant workers vs waiting for migrant workers to navigate their general exclusion from the healthcare system.
Because labour migration and hazardous labour conditions are especially prevalent in low- and middle-income countries (LMICs), there is an urgent and substantial need to assess migrant workers’ health needs and access options to develop and test targeted health interventions specifically designed to reach migrant workers.

## Introduction

Migration is a defining feature of globalisation, with worldwide growing socioeconomic inequalities and shifting workforce demands. According to conservative estimates, the number of international labour migrants has been steadily increasing, reaching 169 million in 2019 (1), with greater estimates of internal labour migration (2). Although many mobile workers are in labour arrangements that generally benefit their income, many are engaged in low-wage jobs associated with health risks (3–6). Moreover, low-wage work is often precarious, i.e., dominated by insecurity, informality, and limited workers’ rights (7–9). Given the multiple disadvantages related to migrant status, especially for irregular international migrants (e.g., possible language barriers, limited social support networks, lack of labour and social protection, poor housing options), low-wage migrant workers are often more vulnerable to exploitation than non-migrant workers and have an increased risk of being trafficked for labour (4, 6, 10). Furthermore, labour migrants are often employed in sectors which are known for exploitative and forced labour conditions (1, 6).

Simultaneously, evidence on the social gradient in health (11, 12) indicates that low-wage work, which is commonly occupied by labour migrants, is associated with poor health outcomes, both directly through harmful work conditions and indirectly because of socioeconomic disadvantages (13–15). Although working conditions may vary geographically and by labour activity, high levels of occupational hazards (e.g., exposure to toxins, frequent accidents, repetitive movements, and extreme temperatures), extensive working hours, insecure employment, and substandard living conditions (including overcrowding and financial insecurity) are widespread (14–20). Indeed, these work conditions are often crudely described as the 3Ds: ‘Dirty’, ‘Difficult’ and ‘Dangerous’. These unhealthy conditions for migrant workers can easily be associated with othering: Social categorisation processes that manifest in social structures, institutions, discourses and language that promote and reinforce group-based inequalities, also faced by other migrant groups (21–23). With a particular emphasis on power asymmetries, othering as an analytical lens points to the intersectionality of different social categories (23) – such as low socioeconomic status or migrant and ethnic minority status in the case of low-wage labour migrants – and their exclusionary, disempowering and marginalising effects (22, 23), which manifest in racism and other forms of social exclusion of labour migrants (7, 14, 17, 24). Consequently, multiple poor health outcomes are associated with the work commonly performed by labour migrants, including conditions that affect their physical (e.g., respiratory, musculoskeletal, dermatological, and infectious diseases, injuries), mental and social health (e.g., violence, substance addiction, isolation, common mental disorders) (4, 5, 8, 14–20, 25–27). In a meta-analysis of data on 7,260 labour migrants, almost half had at least one occupation-related morbidity (10). In addition to general healthcare needs, low-wage migrant workers may face specific or greater health and occupational safety needs that require medical attention than individuals with safer jobs and more health-promoting living and working conditions. Yet, despite their exposure to riskier working and living conditions that may require healthcare, studies repeatedly indicate that low-wage migrant workers often have difficulty accessing healthcare (8, 16, 17, 28, 29).

At the same time as states depend on migrant labour, contemporary health systems generally maintain systemic bordering practices that are agnostic, negligent or, at worst, hostile to mobile populations. That is, health systems are often exclusionary, maintaining institutional bordering that intentionally or inadvertently separates wanted and unwanted service recipients (23, 30). Scholars have noted that many health systems are based on othering as a multidimensional social phenomenon, which helps explain the links between minority status and health inequalities (23). Authors have also highlighted how ‘securitisation’ has served as a vehicle that operationalises power structures (e.g., nationalism, race, gender, class) that may be driven by health concerns and yet negatively affect health access (31). Security structures can set the boundaries that create contested identities, and divisions of who belongs and who is overlooked or actively banished (31, 32). Migrant workers are emblematic of those who are often among those least able to access traditional or mainstream service models (e.g., site-based clinics; health promotion in local languages) and will rely on services that can reach and communicate with them.

Healthcare access has been defined as “the opportunity to have health care needs fulfilled” (33). Levesque et al., propose five access dimensions: approachability, acceptability, availability, affordability and appropriateness of services, which are associated with provider and patient characteristics (33). Many of these features can be found in structural and individual bordering of healthcare access, including questions of ‘us’ and ‘them’ and ‘self’ and ‘other’ identities (34, 35).

Drawing on the five access dimensions, we developed a conceptual framework for this review, which applied commonly reported access barriers (36–38) (Supplement 1). Constraints that often impact populations at large include direct and indirect costs, inadequate insurance coverage, geographical distance, lack of affordable transport, work-related time constraints, and service gaps (14–16, 38). Migrant workers often encounter further access barriers related to their legal status and missing documents (e.g., passport and work permits), language and cultural differences, mobility that hinders the continuity of care, discrimination by health system representatives, and challenges due to being unfamiliar with local care structures and entitlements (4, 26, 36–39). For example, even where documented migrant workers are covered by mandatory healthcare insurance schemes, it is not uncommon for workers to be unaware of their entitlements to care and for medical fees to be higher than for citizens (40). Inequitable healthcare access has been conceptualised as determined by social characteristics and access-enabling resources (e.g., insurance, time, and service availability) rather than need (41). To achieve universal health coverage as envisioned by the United Nations’ Sustainable Development Goals’ target 3.8 (42), and to realise the right to health as a human right (4, 43), health systems need to adapt to the lived realities of low-wage labour migrants, which influence their health needs and form the context of healthcare seeking (Supplement 1).

Several relevant literature reviews have been conducted over the past 20 years, specifically on healthcare services for migrant farmworkers in the USA (26, 44–46). Furthermore, evidence on workplace health promotion programmes for migrant workers across the globe has been compiled, but without including medical services (47). Therefore, despite the need to overall improve healthcare access for low-wage migrant workers (10, 16), knowledge on existing services and their effects remains limited, impeding evidence-informed policies and interventions (4, 8, 26, 46).

To fill this knowledge gap, we reviewed healthcare interventions that targeted migrant workers in sectors associated with low-wage and forced labour. More specifically, we examined how these interventions influenced healthcare access and health-related outcomes (physical and mental health and well-being, service access and uptake, patient satisfaction and acceptability, and cost-effectiveness) (Supplement 1).

## Methods

We did a systematic review following PRISMA guidelines (48) (see Supplement 2 for PRISMA Checklist) and registered a protocol (PROSPERO: CRD42023459360) (49), from which we deviated by narrowing the review’s focus down from low-wage workers in general to low-wage migrant workers.

### Inclusion and exclusion criteria

We included qualitative and quantitative primary studies and reports (published in English or French from 2000 onwards) containing descriptions or evaluations of healthcare interventions exclusively targeting low-wage migrant workers and their families. Full texts needed to detail at least the target population, services provided, and staff composition for inclusion. We aimed to identify interventions that enable migrant workers (and their families) to receive a range of general healthcare services (e.g., general primary medicine, maternal health, dental care, mental health, occupational health services) provided by mobile clinics, clinics on worksites, or other established (or place-based) clinics. To consider a certain level and immediateness of care also allowing for curative elements, only services provided by healthcare professionals (e.g., physicians, nurses, psychiatrists, midwives) were eligible. In addition, we included telehealth interventions to examine approaches for overcoming different access barriers and assuring continuity of care for mobile populations (50, 51). The population of interest is internal and international migrant workers worldwide who are likely to receive low pay under exploitative or otherwise precarious working conditions. We therefore focussed on sectors commonly associated with exploitative work (domestic work, construction, manufacturing, agriculture/forestry/fishing, mining) by drawing on the International Labour Organisation’s 2016 and 2021 Global Estimates of Modern Slavery (6, 52).

We excluded unclear or mixed-income groups, non-migrants, and commercial sex workers (given the comparatively more research on this sector (16, 53–58)) as well as interventions focussing only on specific diseases, vaccination, screening, and emergency care, interventions to increase access to the wider health system, interventions also targeting other patient groups, and health promotion interventions, which have been reviewed elsewhere (17, 47, 59). Inclusion and exclusion criteria were developed within the PICO framework (60) (see Supplement 3).

### Search strategy

We searched Medline, Embase, Web of Science, Global Health, and Global Index Medicus for studies and reports on 9^th^ June 2023 by combining free-text terms and subject headings related to the healthcare interventions AND work conditions AND work sectors of interest (see Supplement 4). To identify further published and grey literature, we simplified the search strategy for searches in Google Search and Google Scholar and hand-searched the bibliographies of all included references. Records were deduplicated (61) and uploaded into Rayyan (62) for duplicate screening. Titles or abstracts had to mention health services for further inclusion. During full text screening, we documented the primary reason for exclusion (Figure 1).

**Figure 1:**
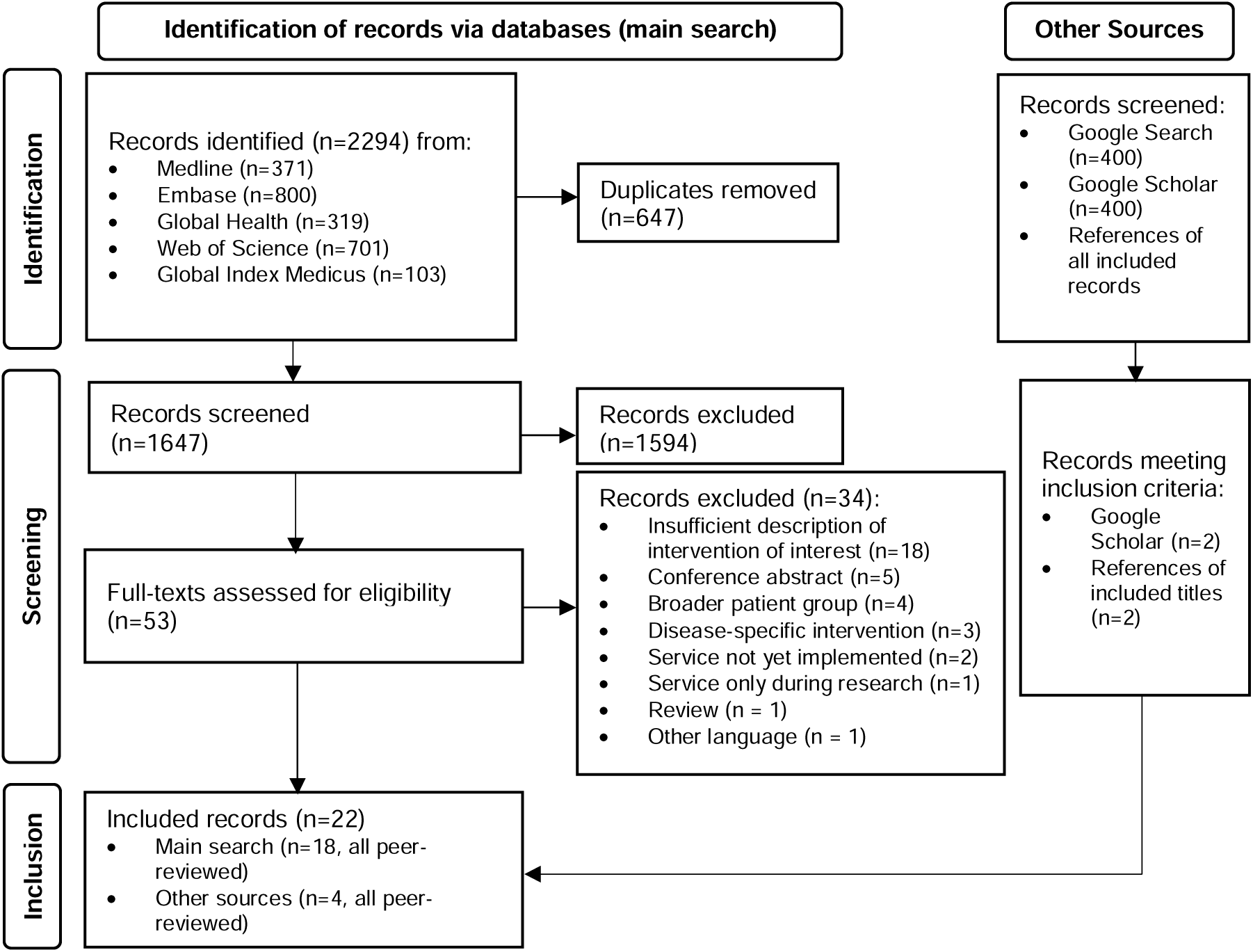
PRISMA flow diagram presenting the selection of references

### Critical appraisal

The quality of those studies examining intervention effects was critically appraised, independently by two reviewers, using JBI Critical Appraisal Tools (63), with scores encompassing low, medium and high study quality. For mixed-methods studies, we appraised the study component (i.e., qualitative or quantitative) reporting relevant outcomes. Discordant appraisals were discussed until an agreement was reached. Quality did not determine inclusion but was considered in the analysis.

### Data extraction

Using a customised form, we extracted general information on the study or report, service characteristics, patient population, context, as well as challenges and facilitators of the intervention. Furthermore, we collected information on how interventions influenced healthcare access within the framework by Levesque et al. (33). For research studies examining intervention effects (on physical and mental health outcomes, patient satisfaction and acceptability, healthcare access and uptake, or cost-effectiveness), we furthermore collected information the relevant outcomes. The first author extracted the data with verification by the second and third authors.

### Data synthesis and analysis

Included interventions were tabulated and ordered by the primary mode of service delivery (i.e., established, mobile, or telehealth service) for sub-group analysis. The first part of the synthesis encompasses all included titles, summarising the interventions’ characteristics and their impact on healthcare access by drawing on the framework by Levesque et al. (33). The second part, limited to a subset of studies, narratively synthesises intervention effects.

## Results

### Characteristics of included studies and reports

Of 2294 records from the databases and further references from other sources (including search of grey literature) we included 22 titles from the academic literature (64–85). Most of them were descriptive reports (66–68, 70, 72, 74, 82) (two relating to the same intervention (66, 67)) or studies not focussing on intervention effects (64, 69, 71, 73, 75, 78, 79, 81, 84). Only six studies (65, 76, 77, 80, 83, 85) examined relevant intervention effects, but were of mixed quality.

### Intervention characteristics

Most of the 21 different healthcare interventions (see overview table in Supplement 5) were implemented in the USA (64, 66–68, 70, 72, 73, 75, 77, 78, 80, 82), followed by the Dominican Republic (69, 79, 83, 85), China (65, 76, 81), Italy (84), Qatar (74), and South Africa (71). Except for two Chinese interventions for internal migrant workers (65, 81), services targeted international migrant workers and their families.

Interventions involving established (or place-based) health facilities (64, 65, 70, 72–75, 77, 81) and mobile clinics (66–69, 71, 78, 79, 82–85) were described by nine and 11 titles respectively. Another two studies reported on telehealth apps (76, 80). Some interventions also combined place-based, outreach, and telehealth services (68, 74, 75). While most titles reported on individual local interventions, a few focussed on the US-wide system of migrant health centres (70, 77) or the subnational occupational health system in the Chinese district Bao’an (65).

Established healthcare interventions were of heterogeneous scales and scope, ranging from a general hospital in an industrial area in Qatar (74) to primary healthcare provided on weekends in established medical centres in the USA (72). Most of these facilities offered primary or occupational health services. The occupational health system in Bao’an was established to complement primary healthcare structures (65). Mobile clinics offered primary healthcare, sometimes also including more specialised services, such as dental, maternal, and paediatric care or physiotherapy (66, 67, 69, 71, 79, 83, 84). The two telehealth interventions were apps for mental health (76) and chronic disease management (80). Health education and other health promotion commonly formed part of interventions (e.g., stretching (64, 82), occupational health and safety measures (64), or patient support groups (71)). A few programmes also addressed wider social determinants of health through food supplementation (79), donated goods (69), or comprehensive social services (68). Healthcare staffing in established facilities ranged from big interdisciplinary and highly specialised teams (74) to nurse-led satellite clinics (75). Mobile clinics were operated by smaller teams of nurses and physicians or by nurses alone (71), but medical specialties were rarely detailed. Some interventions were supported by additional voluntary health professionals, including healthcare students (64, 66, 67, 78, 82), and two were exclusively volunteer-run (69, 72).

Agricultural workers dominated as a target group (16 out of 21 interventions), including all but one intervention in the USA, all in the Dominican Republic, and all mobile clinics (66–73, 75, 77–80, 82–85). Established, non-mobile clinics were also provided in manufacturing (64, 65, 70, 74, 81). The only intervention implemented among (but not exclusively) domestic workers was the mental health app (76). Construction workers were only mentioned once in the context of US migrant health centres, which mainly serve farmworkers (70).

Actors involved in planning and implementing the interventions included private corporations, governmental bodies, academic institutions, and NGOs (iNGOs or local civil society organisations). While no collaboration between NGOs and private actors occurred, all other combinations and individually-led interventions were reported. Established clinics resulted from either governmental (65, 74) or private sector initiatives (81). The governmental occupational health system in Bao’an, e.g., involved factory employers through partial funding and occupational health training (65). Only smaller, semi-permanent services involved NGOs (64, 72), with the exception of federally-qualified health centres in the USA which count as community-based organisations (70, 73, 75, 77). Mobile clinics mostly involved local or iNGOs (69, 83–85), at times with academic (66, 67, 78, 79, 82) and governmental partnerships (71). The two telehealth interventions were developed and implemented by universities (80), in one case supported by community organisations (76).

For about half of the interventions, the source of funding was not discernible. Based on the information available, established health facilities were mainly government-funded (70, 73–75, 77), but the two Chinese industrial clinics were fully or partly paid for by the operating company (65, 81). Mobile clinics were funded by governmental (68), NGO (69, 71, 84, 85), and academic actors (66, 67, 79). Difficulties in acquiring necessary resources, including staff, clinic sites, and financial means, were the most frequently mentioned challenge (65, 66, 71, 75–77), while collaborations with other healthcare providers (70, 71, 74, 75), community organisations (67, 72, 76), and employers (65, 68) were commonly reported as facilitating the interventions.

### Access to healthcare services

The reviewed interventions influenced healthcare access for low-wage migrant workers across Levesque et al.’s (33) five access dimensions (Table 1).

**Table 1:** Healthcare access dimensions addressed by interventions targeting migrant workers

| Access dimension (Levesque et al. (33)) | Barriers in wider health system | Ways in which interventions addressed these barriers |
| --- | --- | --- |
| **Approachability**<br>• Transparency<br>• Outreach<br>• Information<br>• Screening<br>**Ability to perceive**<br>• Health literacy<br>• Health beliefs<br>• Trust<br>• Expectations | Lack of knowledge about services and eligibility | Make services known through outreach activities<br>• with community visits (66, 67)<br>• by involving community support organisations and community health workers (64, 71–73, 76, 79)<br>• by using media channels (radio, television, social media) (64, 76)<br>Support in navigating services<br>• through community health workers (73, 79)<br>• by designing simple care pathways (74)<br>• support with scheduling follow-ups (75, 78)<br>• pointing out healthcare providers in next destination (personal/digital help) (70, 71, 80) |
|  | Health literacy/health beliefs | Provide health education and health screenings (64–69, 72, 73, 75, 78) |
|  | Lack of trust, e.g. due to undocumented legal status | Generate trust through<br>• collaborations with community support groups and community health workers (72, 73)<br>• through long-term (66, 68) and regular engagement (75) with the patient community |
| **Acceptability**<br>• Professional values/norms<br>• Culture<br>• Gender<br>**Ability to seek**<br>• Personal/social values<br>• Culture<br>• Gender<br>• Autonomy | Linguistic/cultural differences | Overcome language barriers with<br>• multilingual healthcare staff (68, 74, 75, 77)<br>• translators and community health workers (64, 66, 73)<br>• multilingual telehealth services (76, 80)<br>Address cultural differences by<br>• employing staff with diverse cultural backgrounds (68, 74) and cultural mediators (84)<br>• providing culturally-sensitive care (e.g., through family-friendly clinic spaces, incorporating patients' concepts of health into the care) (66, 75)<br>Considering stigma of mental health by providing specialised services in polyclinic (74) |
|  | Limited autonomy from employer/<br>Potential job loss | Decrease dependency on employers as a barrier for seeking care through<br>• flexible telehealth services (74, 76)<br>• cooperations with employers (65, 66, 68, 82) |
| **Availability and accommodation**<br>(psychical and timely reachability)<br>• Geographic location<br>• Accommodation<br>• Opening hours | Service gaps (on local or national level) | Increase service coverage through<br>• Providing services (71, 85) or extending existing ones during peak times (66)<br>• employment of community health workers between clinic visits (71) |
|  | Distance, lack of transportation | Overcome transport-related barriers through<br>• the service provision at migrant workers' residency/workplace<br>• mobile and telehealth services |
| Appointment mechanisms<br>**Ability to reach**<br>Living environments<br>Transport<br>Mobility<br>Social support | Time | Harmonise service times with the patients' working hours by<br>providing services during non-traditional hours (64, 66, 68, 77, 78)<br>during working hours in collaboration with employers (82)<br>Reduce the time needed for health seeking through<br>telehealth services (68, 74, 76, 80)<br>proximity of services |
| **Affordability**<br>Direct and opportunity costs<br>**Ability to pay**<br>Income, assets<br>Social capital<br>Insurance | Direct and indirect costs as well as lack of health insurance | Offer free or low-cost services and needed medical supplies, independent of insurance status (exemplified by all interventions) |
|  | Lost wages | Harmonise opening times with working hours of the patients (64, 66, 68, 77, 78) |
| **Appropriateness**<br>(fit between services and client needs, timeliness)<br>Technical and interpersonal quality<br>Adequacy<br>Coordination and continuity<br>**Ability to engage**<br>Empowerment<br>Information<br>Adherence<br>Caregiver support | Inadequate fit between services and needs | Provide services that meet migrant workers' needs<br>with qualified, multidisciplinary healthcare staff trained in needed specialities, such as occupational and mental health (64, 65, 70, 72, 74)<br>by taking into account living and working conditions in the care provision (64, 66)<br>a referral system for more specialised or higher-level care (64, 75, 78, 82)<br>with timely access to care (74) |
|  | Linguistic and cultural differences | Address language and cultural barriers to assure adequate patient-provider communication and the involvement of patient/community representatives (see *acceptability* above) |
|  | Mobility | Assure continuity of care for mobile workers through<br>medical records transfer, transfer letters, or patient-held records (70, 71)<br>handing out higher supplies of medication (71)<br>patient navigation and referral in next destination (70, 71, 80)<br>Provide telehealth services (76, 80) |

To make services known, and thus *approachable*, among target groups, media and personal outreach as well as health education activities and were adopted (64, 66, 67, 76). The involvement of community health workers (CHWs) (71–73, 79), long-term community engagement (66, 68) and regular staff-patient contact (75) reportedly increased trust in services. Navigation of care systems was facilitated through simple pathways (74) or support with follow-ups and referrals (78, 75), including in the next destination of the mobile workers (70, 71, 80).

To increase service *acceptability* for migrant workers, linguistic and cultural differences were addressed, by engaging multilingual staff (68, 74, 75, 77), translators (64, 66), CHWs (73), and cultural mediators (84), or by applying digital tools (76, 80) and incorporating patients’ health beliefs and practices (66, 75). Service acceptability reportedly further increased through telehealth services (making uptake flexible and independent of employer authorisation (74, 76)), employer involvement (65, 66, 68, 82), and by providing mental health services in a general hospital to reduce stigmatisation (74).

As most targeted services were *available* where migrant workers lived or worked or offered telehealth options (68, 74, 76), transport-related barriers (including time and cost) were often circumvented. Clinic times were sometimes harmonised with patients’ working hours by offering weekend or evening services (64, 66, 68, 77, 78). While service availability was overall improved, it varied considerably, from the around-the-clock opening of the Qatari hospital (74) to irregular and intermittent mobile clinic visits and the differing compatibility of clinic and working hours (83, 85).

Most interventions seemed to be *affordable* through low- or no-cost services, since financial constraints were commonly described as impeding access. If detailed, services were mostly free or highly subsidised (64, 65, 67, 69, 71, 72, 74, 84, 85) and accessible independent of insurance status (68).

To ensure *appropriate* type and quality of services, qualified healthcare staff, including with specialisations in mental and occupational health (64, 65, 70, 72, 74) were engaged. Sometimes not all necessary medical specialties were available (69, 83), due to which referrals to other services were made (64, 75, 78, 82). However, other transport- and cost-related barriers could continue to impede access into wider care structures (64). In some cases, migrant workers’ living situation (66) and mobility were considered when providing and planning treatments: Continuity of care for mobile workers was sought through telehealth interventions (76, 80), virtual care management in the USA (offering navigation support, transfer of medical records, and referrals) (70), and patient-held medical records, higher medication supplies, and transfer letters in South Africa (71).

### Intervention effects

The subset of six studies with widely ranging participant numbers and mixed quality provide scattered evidence on intervention effects (Table 2).

**Table 2:**
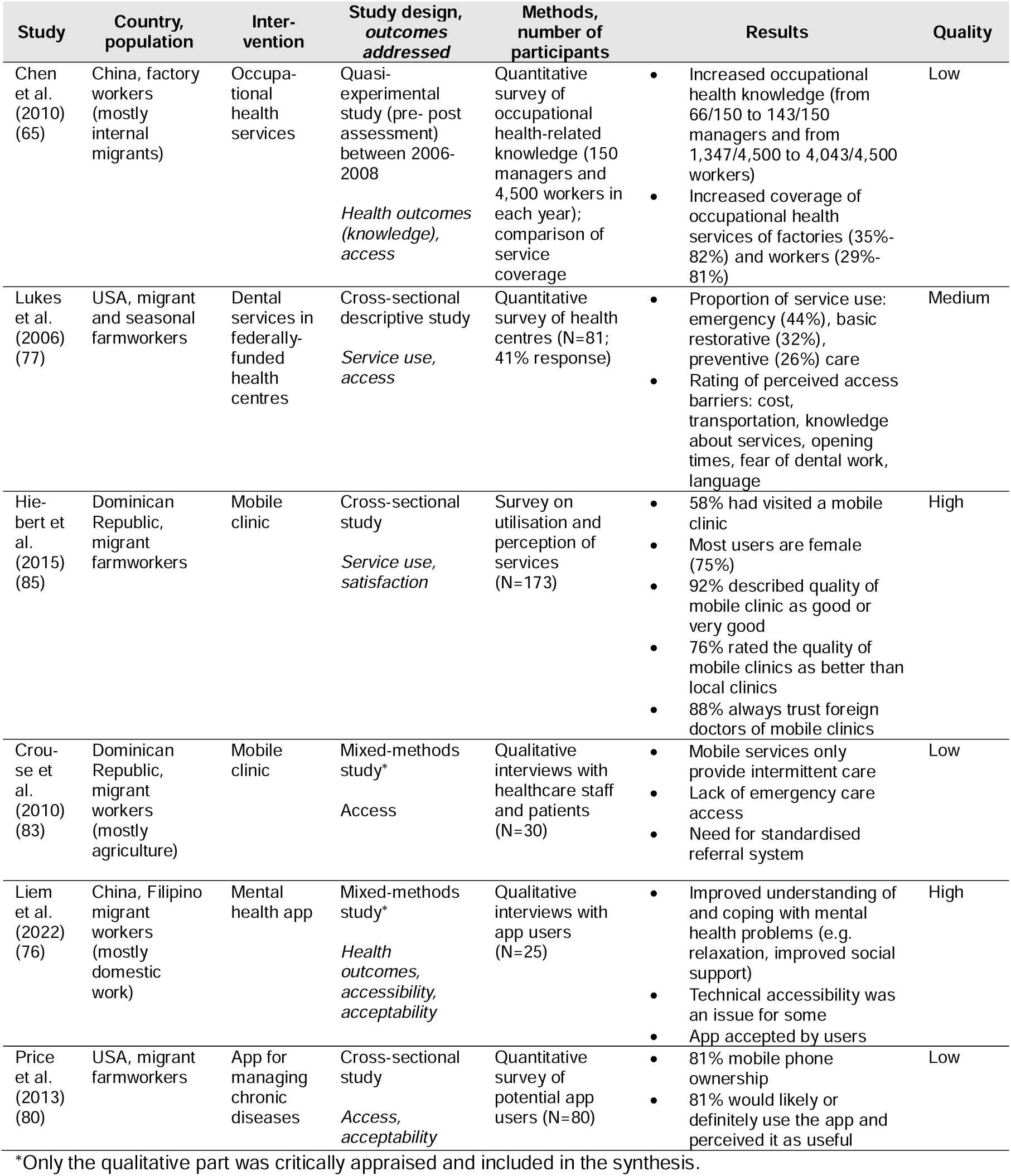
Overview of studies evaluating intervention effects (N=6)

| Study | Country, population | Intervention | Study design, outcomes addressed | Methods, number of participants | Results | Quality |
| --- | --- | --- | --- | --- | --- | --- |
| Chen et al. (2010) (65) | China, factory workers (mostly internal migrants) | Occupational health services | Quasi-experimental study (pre- post assessment) between 2006-2008<br>*Health outcomes (knowledge), access* | Quantitative survey of occupational health-related knowledge (150 managers and 4,500 workers in each year); comparison of service coverage | Increased occupational health knowledge (from 66/150 to 143/150 managers and from 1,347/4,500 to 4,043/4,500 workers)<br>Increased coverage of occupational health services of factories (35%-82%) and workers (29%-81%) | Low |
| Lukes et al. (2006) (77) | USA, migrant and seasonal farmworkers | Dental services in federally-funded health centres | Cross-sectional descriptive study<br>*Service use, access* | Quantitative survey of health centres (N=81; 41% response) | Proportion of service use: emergency (44%), basic restorative (32%), preventive (26%) care<br>Rating of perceived access barriers: cost, transportation, knowledge about services, opening times, fear of dental work, language | Medium |
| Hiebert et al. (2015) (85) | Dominican Republic, migrant farmworkers | Mobile clinic | Cross-sectional study<br>*Service use, satisfaction* | Survey on utilisation and perception of services (N=173) | 58% had visited a mobile clinic<br>Most users are female (75%)<br>92% described quality of mobile clinic as good or very good<br>76% rated the quality of mobile clinics as better than local clinics<br>88% always trust foreign doctors of mobile clinics | High |
| Crouse et al. (2010) (83) | Dominican Republic, migrant workers (mostly agriculture) | Mobile clinic | Mixed-methods study*<br>Access | Qualitative interviews with healthcare staff and patients (N=30) | Mobile services only provide intermittent care<br>Lack of emergency care access<br>Need for standardised referral system | Low |
| Liem et al. (2022) (76) | China, Filipino migrant workers (mostly domestic work) | Mental health app | Mixed-methods study*<br>*Health outcomes, accessibility, acceptability* | Qualitative interviews with app users (N=25) | Improved understanding of and coping with mental health problems (e.g. relaxation, improved social support)<br>Technical accessibility was an issue for some<br>App accepted by users | High |
| Price et al. (2013) (80) | USA, migrant farmworkers | App for managing chronic diseases | Cross-sectional study<br>*Access, acceptability* | Quantitative survey of potential app users (N=80) | 81% mobile phone ownership<br>81% would likely or definitely use the app and perceived it as useful | Low |

\*Only the qualitative part was critically appraised and included in the synthesis.

Two studies examined health-related outcomes, pointing to improved mental health literacy and well-being through a mental health app for Filipino migrant workers (based on qualitative interviews) (76) and increased occupational health-related knowledge of Chinese factory employers after two years of occupational health services and training (based on survey data) (65).

Healthcare access and uptake were examined in different ways. The coverage of the occupational health system in Bao’an increased from 610,000 to 1.9 million workers and from 35% to 82% of factories between 2006 and 2008 (65). Lukes et al. surveyed health centres across the USA providing dental services to migrant farmworkers. Service use was dominated by emergency care (44%), while restorative (32%) and preventative (26%) services accounted for fewer visits than aspired. Health centre representatives ranked cost and transport (same ranking), insufficient knowledge of services, limited clinic hours, and language barriers as the most common access barriers (77). Hiebert et al. (85) found that young adults and males in agricultural communities in the Dominican Republic visited mobile clinics less frequently than women and older people. Based on qualitative interviews with farmworkers and healthcare providers in the Dominican Republic, Crouse et al. (83) reported lacking emergency care access between periodic mobile clinic visits and the need for a standardised referral system for higher level care. The two telehealth studies evaluated accessibility in terms of mobile phone ownership, which was found to be high (81% in 2011-2012) among migrant farmworkers in the USA (80), and technical accessibility while using the app, where Filipino migrant workers encountered different technical issues (76).

Three studies evaluated patient satisfaction and acceptability, indicating positive effects. Surveys yielded superior patient assessments of mobile clinics compared to local services in the Dominican Republic (85) and high levels of willingness to use the app for chronic disease management among farmworkers in the USA reported they would likely or definitely use (80). Liem et al. (76) concluded from qualitative interviews that the mental health app for overseas Filipino workers was well-accepted.

## Discussion

Migrant workers comprise one of the most important cohorts in the world’s basic production and service sectors. They are also often the individuals who are exposed to the greatest health risks and most substantial barriers to healthcare based on multidimensional othering and systemic bordering of labour and social protections and health services. This review identified 21 health-related interventions for migrant workers in six countries that attempt to overcome systemic borders. These interventions included diverse models of care, including mobile clinics, established healthcare facilities and telehealth interventions provided by governmental, NGO, academic, and private actors. Ultimately, however, most documented interventions targeted farmworkers and were based in the USA, while none were identified in low and lower-middle income countries.

### Intervention effects

The extent to which the interventions influenced workers’ access and uptake, their health, patient satisfaction, and acceptability or were cost-effective remains largely unknown due to limited evaluative evidence. This finding echoes previous remarks about the need for more intervention research and evaluations on health services for low-wage and migrant workers (4, 26, 46, 85), and mobile clinics, in general (86, 87). However, the absence of evaluations might also be an artefact of the database-focussed search strategy, since programme evaluations are not always published in academic forums (46, 87).

### Healthcare access

The interventions adopted various strategies to address commonly reported access barriers that exclude low-wage and migrant workers from health systems. Results indicate that financial barriers were overcome almost universally through low-cost or free care. While the availability of healthcare services was generally improved, findings also indicate remaining service gaps due to intermittent outreach visits or the incompatibility of service times with patients’ working hours. Furthermore, mobile or smaller place-based clinics only offered a limited range of services, often contingent on individual staff members, which potentially decreases the appropriateness of care, i.e., the fit between services and patient needs (33). Many publications mentioned that interventions addressed linguistic and cultural differences. However, it often remained unclear how (and whether) this was achieved and perceived by patients or whether othering was (unintentionally) reinforced by reproducing specific social categories. Truly non-discriminatory and patient-centred care avoids cultural essentialism and the reduction of “culture” to language. Healthcare services for indigenous communities, often based on community engagement, offer valuable examples for effectively making services culturally-sensitive (88). Services were also made more appropriate for mobile workers by assuring continuity of care, e.g., through transferred or patient-held medical records. The US Health Network, a virtual case management with links to 120 countries (89), exemplifies cross-border care that benefits mobile patients as a whole. While the different ways in which the reviewed interventions facilitated healthcare access may offer valuable examples for overcoming the multidimensional access barriers commonly faced by low-wage migrant workers, the overall accessibility of services remains unclear.

Based on the review findings, the potential of telehealth for this mobile population seems to be relatively untapped. Telehealth services generally show high effectiveness (90), which can lower access barriers related to service gaps, transport, language, and time, and decrease dependency on employer consent for care seeking. However, telehealth can also reconfigure barriers (91). For example, the Non-Resident Nepali Association organised multidisciplinary telemedicine services during the COVID-19 pandemic to connect Nepalis based abroad with health professionals through various digital technologies (e.g., email, telephone, video calls). Insufficient transborder regulations for providing medical consultations and prescriptions, digital gaps, and low literacy levels of some patients posed challenges (92). Technology and literacy barriers have also been reported for other populations using telehealth services, e.g., racial and ethnic minority groups (51, 91). Telehealth interventions may thus also increase inequities in access for migrant workers and therefore need careful planning.

Engagement of patient and community members figured across the five access dimensions in this review. In particular, CHWs linked patients and services, making services more approachable through information and trust-building, lowering linguistic and cultural barriers, and improving availability through basic healthcare. A recent review by the World Health Organisation concluded that CHWs have “enormous potential to extend health care services to vulnerable populations”, including through curative services (93). The sustainability and effectiveness of CHW programmes was improved by their embeddedness in national health systems and communities as well as appropriate training and support of CHWs (93).

### Integration with wider health systems

This review raises the question of service integration into wider care structures, which varied across the reviewed interventions. While the Qatari industrial hospital and occupational health system in Bao’an complemented established primary healthcare structures and a country-wide network of migrant health clinics exists in the USA, formalised links to other healthcare services were lacking for some of the identified mobile clinics or barriers (e.g., related to transport and costs) or were dysfunctional due to persisting access barriers.

For healthcare to be appropriate, services must meet needs (33), which requires referral options for more complex needs, as stressed by the International Committee of the Red Cross mobile clinic directives (94). This poses particular challenges where health systems are overburdened. Non-governmental and private corporate activities can fill resulting service gaps but are often of limited sustainability and scope and may trigger service fragmentation (71, 95, 96). Thus, a reliance on non-governmental actors can undermine overall health system strengthening (96). Furthermore, employer-provided healthcare might be unacceptable for workers who fear negative repercussions from disclosing ill-health (14).

Ultimately, national governments are responsible for population health (43) but these are also the same entities that intentionally or neglectfully structure health systems that exclude or omit migrants. Indeed, a recent UK study on healthcare and education structures highlighted the securitization of these basic services by requiring data-sharing to advance the UK Home Office immigration agenda (34). Similarly, securitization of health in LMICs, such as Malaysia, intensified during the COVID-19 pandemic, deterring undocumented migrants from accessing essential healthcare services, which hindered both preventive and curative efforts (32).

To achieve advancements towards health equity, health systems must offer diversity-sensitive services that make appropriate efforts to include migrant workers, independent of their immigration status (4, 29). Service delivery must consider the multidimensional bordering that excludes workers by integrating interventions that are sensitive to diverse needs, especially of full-time workers, into national health systems, while avoiding parallel and unsustainable structures. Diversity-sensitive systems will benefit from a migrant patient-centred understanding of healthcare and access priorities. For instance, undocumented migrants in Italy have legal access to health services (4), but the reviewed mobile clinic for migrant farmworkers in Italy (84) illustrates that these entitlements cannot be equated with an actual opportunity for access. Therefore, until health systems provide equitable access to this population, targeted interventions have to bridge prevailing gaps but should not function in isolation. In fact, collaborations with other healthcare providers, NGOs, and employers were identified as a major facilitator among the reviewed interventions. This is congruent with findings from a related review, attributing mobile clinic sustainability to long-term involvement of different organisations, including academic and community partners (46). Thus, vertical approaches specifically targeting the needs of low-wage labour migrants are needed but should converge with horizontal efforts that aim to improve the accessibility of health systems more broadly.

### Living and working conditions

While beyond the focus of the present study, it needs to be acknowledged that healthcare is only one, and not necessarily the most impactful, determinant of health on a population level (97, 98). Thus, in addition to providing accessible healthcare, other multilevel and multisectoral approaches are needed for improving the health of low-wage workers (14, 15). Importantly, the living and working conditions that also influence healthcare needs and the possibilities for healthcare access (Supplement 1) need to be assessed and addressed. Health promotion interventions (17, 47, 59, 99) may contribute to general health protection, combined with structural level shifts. For example, in line with the International Labour Organisation’s Decent Work Agenda (100) health promotion for migrant workers would include humane immigration laws, workplace health and safety regulations, paid sick leave, adequate social protection and living wages (14, 15). For instance, findings from a natural experiment indicate that the introduction of minimum wages in the United Kingdom in 1999 significantly improved low-wage workers’ mental health (101).

### Limitations

When interpreting these results, the limitations and characteristics of the identified body of evidence have to be considered. There was a strong focus on the agricultural sector, migrant workers, and, geographically, the USA – reflecting bibliometric findings (102). Included titles contained varying levels of relevant information, which was mostly descriptive. Theoretical underpinnings of the interventions were overall lacking. The sparse evaluative evidence was mostly of limited quality.

The perspective offered by the present review is limited by its methodological approach, including the language constraints and the briefness of the grey literature search. Furthermore, while the search terms the work sectors were selected carefully, they do not cover the entire global population of low-wage labour migrants, who are engaged in a wide range of activities. Relevant interventions offering valuable insights might also have been missed by excluding services also targeting other patient groups or interventions that aim to facilitate access to wider care structures.

### Implications

This review has important implications for overcoming othering and healthcare bordering practices, and improving policies for inclusive systems. For healthcare decision-makers, findings indicate common access barriers and forms of exclusion, and suggest strategies designed to respond to migrant workers’ needs, which can be adopted by health systems. Achieving inclusive healthcare and move towards greater health equity in responding to social determinants of health will benefit from multisectoral collaborations, e.g., between healthcare providers, governmental agencies, employers, and, importantly, the patient community.

Budget allocations are central to making health systems accessible and effective for migrant workers and similarly excluded population groups. Moreover, the medical profession will benefit from including training on migrant-inclusive services into medical curricula and clinical practice. For example, occupational safety and health, plus diversity-sensitive service essentials should form part of medical school curricula and migrant-aware clinical intake processes (103). Moreover, inclusive health strategies will subsidise care for uninsured and undocumented patients (29, 103), including adequate insurance coverage and making health systems more “migration-aware” (104). Internationally, regulations for cross-border healthcare provision, including prescriptions, need to be further established, perhaps by exploring the potential of telehealth strategies (92). As noted, healthcare policies cannot be undermined by exclusionary labour, immigration, and social policies.

Apart from the general need for more research on low-wage and migrant workers’ health (102), more studies on targeted interventions are needed – preferably with longitudinal mixed-method designs capturing longer-term effects, including on equity in access, cost-effectiveness, programme sustainability, and patient perspectives. In particular, the potential of telehealth services for this mobile population should be further examined. In parallel, more extensive reviews of literature available outside of academic forums should be undertaken and past evaluations should be made more widely available. In addition to targeted interventions, the research focus should also be directed to measures aiming to facilitate migrant workers’ access into the wider health system (e.g., through CHWs or insurance schemes).

## Conclusion

Low-wage migrant workers are a heterogeneous population who sustain numerous crucial labour sectors, yet they often encounter multiple health risks and exclusion from healthcare. Given the global prevalence of labour migration (1, 2), health equity via universal health coverage can only be achieved if we meet the healthcare needs of migrant workers. Healthcare, while integral, can only be part of a strategy to protect the health of these not invisible but often overlooked workers.

## Supporting information

Supplement1_Framework

Supplement2_PRISMA

Supplement3_PICO

Supplement4_SearchStrategies

Supplement5_StudyOverview

## Data Availability

This review is based on published literature only.

## Declarations

### Ethics approval and consent to participate

Not applicable

### Consent for publication

Not applicable

### Availability of data and materials

This review is based on published literature only.

## Authors’ contributions

CZ proposed the initial idea for the review. ER drafted the search strategy, which was reviewed with the help of CZ, KL, SH and a health librarian. ER performed all stages of the screening process and quality appraisal. KL and RCL duplicated part of the screening process and quality appraisal. ER performed the data synthesis. Results were discussed with all authors. ER drafted the manuscript, which all authors reviewed for intellectual content. All authors read and approved the final manuscript.

## Acknowledgements

We thank Russell Burke from the library team at the London School of Hygiene & Tropical Medicine for his support in the development of the search strategy. Our thanks furthermore go to Nicola Pocock for methodological advice.

## Abbreviations

CHW: Community health worker
COVID-19: Corona virus disease of 2019
ICRC: International Committee of the Red Cross
iNGO: International non-governmental organisation
LMICs: Low- and middle-income countries
LSHTM: London School of Hygiene & Tropical Medicine
JBI: Joanna Briggs Institute
NGO: non-governmental organisation
PHC: Primary healthcare
PICO: Population, Intervention, Comparator, Outcome
PRISMA: Preferred Reporting Items for Systematic Reviews
PROSPERO: International Prospective Register of Systematic Reviews

