## Supplement1_Framework for "Healthcare interventions for low-wage migrant workers: A systematic review"

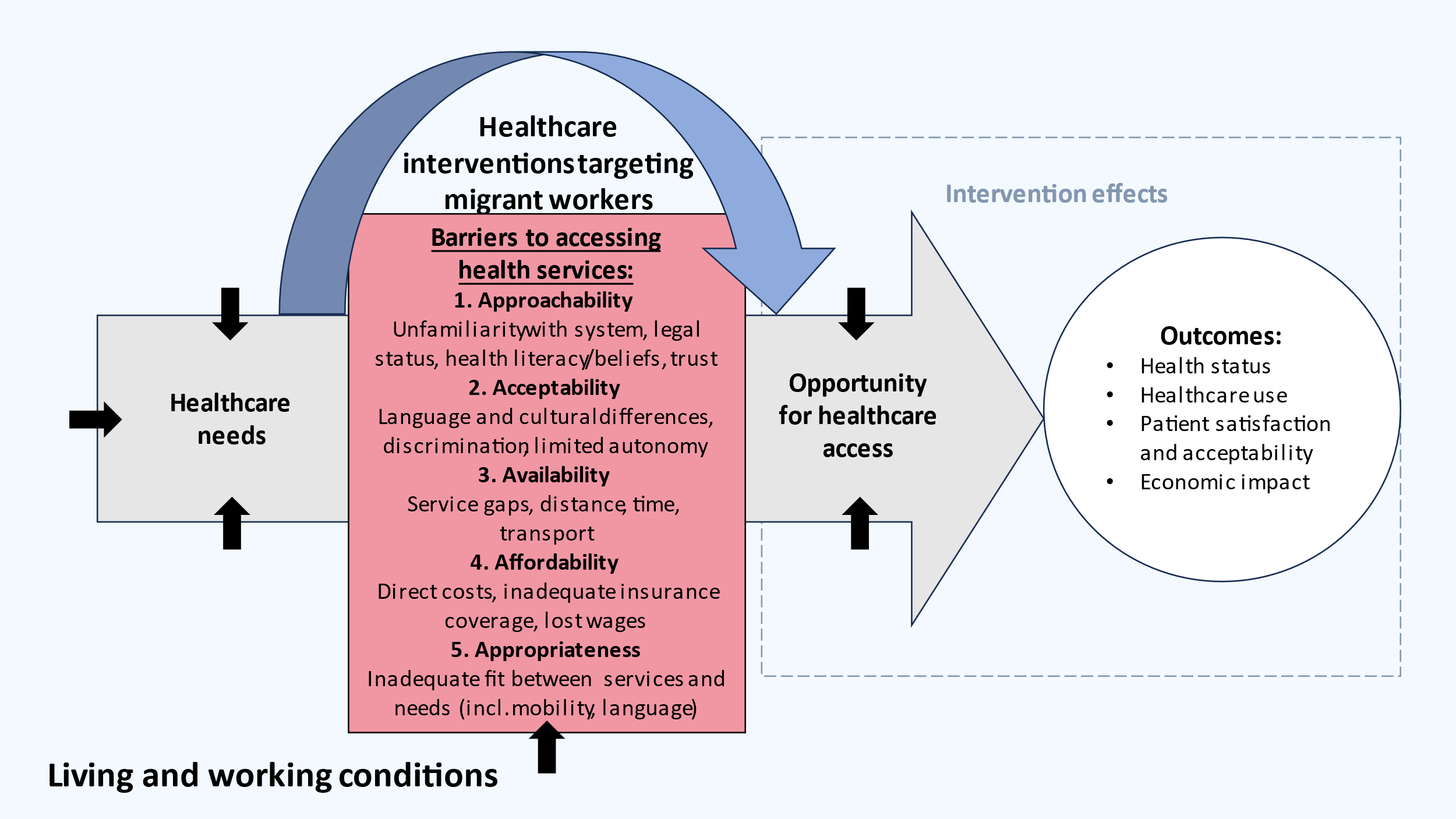

**Supplement 1:** Conceptual framework for the present review of healthcare interventions for low-wage migrant workers
