## Supplement3_PICO for "Healthcare interventions for low-wage migrant workers: A systematic review"

### **Supplement 3:** Inclusion criteria developed in PICO(S) framework

|  | **Inclusion** | **Exclusion** |
| --- | --- | --- |
| **Population** | Low-wage labour migrants, who work e.g. (but not exclusively) in domestic work, construction, mining, manufacturing agriculture/forestry/fishing; no geographical limitation | Mixed patient populations of different income groups or population where it is unclear whether it is restricted to work sectors that are associated with low-wage work; sex workers are excluded |
| **Intervention** | - Mobile clinics or - On-site or otherwise established healthcare services   which specifically target migrant workers and their families and provide a range of general health care services that also include curative elements (e.g. general, maternal, dental, mental, and occupational health services) and are provided by professional health care staff (e.g. nurses, doctors, midwives, and psychiatrists)   - or telehealth interventions that have been implemented among low-wage workers | - disease-specific interventions (e.g. TB, HIV, malaria, screening for specific diseases) - emergency care without primary health care - One-off immunisation/ disease control interventions (e.g. during disease outbreaks) - Interventions consisting only of health promotion, including tobacco control, injury prevention, contraception, or purely health education - Policies that aim to decrease barriers for health seeking for migrant workers - Single health care/ counselling sessions that were offered as part of a research study - services specifically targeting sex workers - Mandatory health screenings for migrant workers upon entry into the host country - Interventions that aim to increase uptake of health care within the wider health system (which is not only targeting low-wage workers) - Services exclusively provided by lay/ community health care workers - Interventions that have not yet been implemented or piloted |
| **Outcome** | For evaluation studies: e.g. physical and mental health outcomes, patient satisfaction, health care access, cost-effectiveness | No exclusion based on outcomes (as evaluation is not a prerequisite for inclusion of a study) |
| **Study design and content** | Qualitative and quantitative primary studies or reports that **describe** the characteristics of the healthcare services (e.g. services provided, staff members, funding, challenges, facilitators) **and/or** **evaluate** their effects (i.e. descriptive as well as evaluative publications are included) | - Studies/reports that only briefly mention health care interventions but do not give a minimum level of detail on their characteristics (i.e. mode of delivery, patient population, services provided, staff members) or evaluate their effects - Conference proceedings, study protocols - Literature reviews will not be included in the synthesis but considered in discussion/background |
