## Supplement4_SearchStrategies for "Healthcare interventions for low-wage migrant workers: A systematic review"

| **#** | **Query Medline(R) ALL** | **Results from 9 Jun 2023** |
| --- | --- | --- |
| 1 | hospitals/ or exp hospitals, community/ or exp community health services/ or exp "delivery of health care"/ or exp Primary Health Care/ or exp Ambulatory care facilities/ or exp mobile health units/ or exp Telemedicine/ | 1,662,110 |
| 2 | (hospital* or clinic* or medi-point* or infirmary or infirmaries or ambulatory care or primary care or ((health or health care or care or medical or ambulatory) adj1 (post* or cent* or station* or unit* or service* or point* or facility or facilities)) or (outreach adj (care or health or clinic* or consultation*)) or community health or outpatient or telemedicine or tele-medicine or teleconsult* or tele-consult* or telepsychiatry or tele-psychiatry or telepsychology or tele-psychology or telehealth or tele-health or e-health or electronic health or m-health or mobile health).mp. [mp=title, book title, abstract, original title, name of substance word, subject heading word, floating sub-heading word, keyword heading word, organism supplementary concept word, protocol supplementary concept word, rare disease supplementary concept word, unique identifier, synonyms, population supplementary concept word, anatomy supplementary concept word] | 7,861,096 |
| 3 | exp Human Trafficking/ or exp Enslavement/ or exp Child Labor/ | 899 |
| 4 | (modern slavery or ((labo?r or work or job* or occupation*) adj3 (forced or precarious or exploit* or informal or unprotected or bonded)) or ((labo?r* or work*) adj1 (migrant* or migrat*)) or child labo?r or low-wage or low pay or low paid or seasonal work* or sweat shop* or day labo?r* or human traffick*).mp. [mp=title, book title, abstract, original title, name of substance word, subject heading word, floating sub-heading word, keyword heading word, organism supplementary concept word, protocol supplementary concept word, rare disease supplementary concept word, unique identifier, synonyms, population supplementary concept word, anatomy supplementary concept word] | 8,852 |
| 5 | exp Agriculture/ or exp Manufacturing Industry/ or exp "Extraction and Processing Industry"/ or exp Construction Industry/ or exp Tobacco Industry/ or exp Meat-Packing Industry/ or exp Textile Industry/ or exp Food Industry/ or exp Coal Industry/ or exp Food-Processing Industry/ | 413,963 |
| 6 | (domestic work* or domestic labo?r* or construction or industry or industries or factory or factories or fabricat* or manufactur* or garment* or textile* or food processing or agricultur* or farm work* or farmwork* or (plantation* adj4 work*) or fishing or sea slavery or seafar* or forestry or mining or quarry* or (extract* adj4 (gas or petroleum or oil))).mp. [mp=title, book title, abstract, original title, name of substance word, subject heading word, floating sub-heading word, keyword heading word, organism supplementary concept word, protocol supplementary concept word, rare disease supplementary concept word, unique identifier, synonyms, population supplementary concept word, anatomy supplementary concept word] | 1,023,559 |
| 7 | (1 or 2) and (3 or 4) and (5 or 6) | 627 |
| 8 | limit 7 to yr="2000 -Current" | 371 |

| **#** | **Embase** | **Results from 9 Jun 2023** |
| --- | --- | --- |
| 1 | exp hospital/ or exp community hospital/ or exp rural hospital/ or exp field hospital/ or primary health care/ or exp telemedicine/ or exp health care delivery/ | 5013128 |
| 2 | (hospital* or clinic* or medi-point* or infirmary or infirmaries or ambulatory care or primary care or ((health or health care or care or medical or ambulatory) adj1 (post* or cent* or station* or unit* or service* or point* or facility or facilities)) or (outreach adj (care or health or clinic* or consultation*)) or community health or outpatient or telemedicine or tele-medicine or teleconsult* or tele-consult* or telepsychiatry or tele-psychiatry or telepsychology or tele-psychology or telehealth or tele-health or e-health or electronic health or m-health or mobile health).mp. [mp=title, book title, abstract, original title, name of substance word, subject heading word, floating sub-heading word, keyword heading word, organism supplementary concept word, protocol supplementary concept word, rare disease supplementary concept word, unique identifier, synonyms, population supplementary concept word, anatomy supplementary concept word] | 15196881 |
| 3 | exp human trafficking/ or exp slavery/ or exp child labor/ or migrant worker/ | 3858 |
| 4 | (modern slavery or ((labo?r or work or job* or occupation*) adj3 (forced or precarious or exploit* or informal or unprotected or bonded)) or ((labo?r* or work*) adj1 (migrant* or migrat*)) or child labo?r or low-wage or low pay or low paid or seasonal work* or sweat shop* or day labo?r* or human traffick*).mp. [mp=title, book title, abstract, original title, name of substance word, subject heading word, floating sub-heading word, keyword heading word, organism supplementary concept word, protocol supplementary concept word, rare disease supplementary concept word, unique identifier, synonyms, population supplementary concept word, anatomy supplementary concept word] | 10303 |
| 5 | exp agriculture/ or exp fishing/ or exp manufacturing/ or exp construction worker/ or exp construction work/ or exp factory/ or exp factory worker/ or exp meat industry/ or exp textile industry/ or exp food industry/ or exp manufacturing industry/ or exp building industry/ or exp tobacco industry/ or exp industry/ or exp mining/ | 690531 |
| 6 | (domestic work* or domestic labo?r* or construction or industry or industries or factory or factories or fabricat* or manufactur* or garment* or textile* or food processing or agricultur* or farm work* or farmwork* or (plantation* adj4 work*) or fishing or sea slavery or seafar* or forestry or mining or quarry* or (extract* adj4 (gas or petroleum or oil))).mp. [mp=title, book title, abstract, original title, name of substance word, subject heading word, floating sub-heading word, keyword heading word, organism supplementary concept word, protocol supplementary concept word, rare disease supplementary concept word, unique identifier, synonyms, population supplementary concept word, anatomy supplementary concept word] | 1227740 |
| 7 | (1 or 2) and (3 or 4) and (5 or 6) | 895 |
| 8 | Limit 7 to yr=”2000 -Current” | 800 |

| **#** | **Global Health** | **Results from 9 Jun 2023** |
| --- | --- | --- |
| 1 | hospitals/ or exp health centres/ or exp health services/ or exp medical services/ or exp primary health care/ or community health/ or exp community health services/ or exp telemedicine/ | 218595 |
| 2 | (hospital* or clinic* or medi-point* or infirmary or infirmaries or ambulatory care or "primary care" or ((health or "health care" or care or medical or ambulatory) adj1 (post* or cent* or station* or unit* or service* or point* or facility or facilities)) or (outreach adj (care or health or clinic* or consultation*)) or "community health" or outpatient or telemedicine or tele-medicine or teleconsult* or tele-consult* or telepsychiatry or tele-psychiatry or telepsychology or tele-psychology or telehealth or tele-health or e-health or electronic health or m-health or mobile health) | 1309779 |
| 3 | exp child labour/ or exp seasonal labour/ or exp bonded labour/ or exp forced labour/ or exp migrant labour/ | 1871 |
| 4 | ("modern slavery" or ((labo?r or work* or job* or occupation*) adj3 (forced or precarious or exploit* or forced or informal or unprotected or bonded)) or ((labo?r* or work*) adj1 (migrant* or migrat*)) or "child labo?r" or low-wage or "low pay" or "low paid" or "seasonal work*" or "sweat shop*" or "day labo?r*" or "human traffick*") | 5125 |
| 5 | exp manufacture/ or exp rubber industry/ or exp "oil and gas industry"/ or exp tea industry/ or exp textile industry/ or exp coffee industry/ or exp "meat and livestock industry"/ or exp cotton industry/ or exp fish industry/ or exp tobacco industry/ or exp cocoa industry/ or exp building industry/ or exp food industry/ or exp agriculture/ or exp farm workers/ or exp fishing/ or exp mining/ | 37293 |
| 6 | ("domestic work*" or "domestic labo?r*" or construction or industry or industries or factory or factories or fabricat* or manufactur* or garment* or textile* or "food processing" or agricultur* or "farm work*" or farmwork* or (plantation* adj4 work*) or fishing or "sea slavery" or seafar* or forestry or mining or quarry* or (extract* adj4 (gas or petroleum or oil))) | 394,901 |
| 7 | (1 or 2) and (3 or 4) and (5 or 6) | 409 |
| 8 | Limit 7 to yr=”2000 -Current” | 319 |

| **#** | **Web of Science** | **^9th^ June** |
| --- | --- | --- |
|  | TS=(hospital* or clinic* or medi-point* or infirmary or infirmaries or "ambulatory care" or "primary care" or ((health or "health care" or care or medical or ambulatory) NEAR/1 (post* or cent* or station* or unit* or service* or point* or facility or facilities)) or (outreach NEAR/1 (care or health or clinic* or consultation*)) or "community health" or outpatient or telemedicine or tele-medicine or teleconsult* or tele-consult* or telepsychiatry or tele-psychiatry or telepsychology or tele-psychology or telehealth or tele-health or e-health or "electronic health" or m-health or "mobile health")  AND  TS=("modern slavery" or ((labo$r or work or job* or occupation*) NEAR/3 (forced or precarious or exploit* or informal or unprotected or bonded)) or ((labo$r or work*) NEAR/1 (migrant* or migrat*)) or "child labo$r*" or low-wage or "low pay" or "low paid" or "seasonal work*" or "sweat shop*" or "day labo$r*" or "human traffick*")  AND  TS=("domestic work*" or "domestic labo$r" or construction or industry or industries or factory or factories or fabricat* or manufactur* or garment* or textile* or "food processing" or agricultur* or "farm work*" or farmwork* or (plantation* NEAR/4 work*) or fishing or "sea slavery" or seafar* or forestry or mining or quarry* or (extract* NEAR/4 (gas or petroleum or oil))) | 746 |
|  | Timespan: 2000-01-01 to 2023-06-09 (Index Date) | 701 |

| **Global Index Medicus** | **Results from 9 Jun 2023** |
| --- | --- |
| tw:((mh:(hospitals OR telemedicine OR "hospitals, community" OR "community health services" OR "delivery of health care" OR "primary health care" OR "ambulatory care facilities" OR "mobile health units" OR telemedicine)) OR (tw:(hospital* OR clinic* OR medi-point* OR infirmary OR infirmaries OR "ambulatory care" OR primary care OR "health post*" OR "health cent*" OR "health station*" OR "health unit*" OR "health service*" OR "health point*" OR "health facility" OR "health facilities" OR "health care post*" OR "health care cent*" OR "health care station*" OR "health care unit*" OR "health care service*" OR "health care point*" OR "health care facility" OR "health care facilities" OR "care post*" OR "care cent*" OR "care station*" OR "care unit*" OR "care service*" OR "care point*" OR "care facility" OR "care facilities" OR "medical post*" OR "medical cent*" OR "medical station*" OR "medical unit*" OR "medical service*" OR "medical facility" OR "medical facilities" OR "ambulatory post*" OR "ambulatory cent*" OR "ambulatory station*" OR "ambulatory unit*" OR "ambulatory service*" OR "ambulatory point*" OR "ambulatory facility" OR "ambulatory facilities" OR "outreach care" OR "outreach health" OR "outreach clinic*" OR "outreach consultation*" OR "community health" OR outpatient OR telemedicine OR tele-medicine OR teleconsult* OR tele-consult* OR telepsychiatry OR tele-psychiatry OR telepsychology OR tele-psychology OR telehealth OR tele-health OR e-health OR "electronic health" OR m-health OR "mobile health")))) AND (tw:((mh:("Human Trafficking" OR "Enslavement" OR "Child Labor")) OR (tw:("modern slavery" OR "forced labor" OR "forced labour" OR "exploitative labour" OR "exploitative labor" OR "precarious work*" OR "exploitative labour" OR "exploitative labor" OR "labour exploitation" OR "labor exploitation" OR "informal work" OR "informal sector*" OR "unprotected job*" OR "bonded labour*" OR "bonded labor*" OR "labour migrant*" OR "labour migra*" OR "labor migrant*" OR "labor migra*" OR "migrant work*" OR "child labour*" OR "child labor*" OR low-wage OR "low pay" OR "low paid" OR "seasonal work*" OR "sweat shop*" OR "day labour*" OR "day labor*" OR "human traffick*")))) AND (tw:((mh:(agriculture OR "Manufacturing Industry" OR "Extraction and Processing Industry" OR "Construction Industry" OR "Tobacco Industry" OR "Meat-Packing Industry" OR "Textile Industry" OR "Food Industry" OR "Coal Industry" OR "Food-Processing Industry")) OR (tw:("domestic work*" OR "domestic labour*" OR "domestic labor*" OR construction OR industry OR industries OR factory OR factories OR fabricat* OR manufactur* OR garment* OR textile* OR "food processing" OR agricultur* OR "farm work*" OR farmwork* OR "plantation work*" OR fishing OR "sea slavery" OR seafar* OR forestry OR mining OR quarry*))) AND (year_cluster:[2000 TO 2023]) | 105 |

**Grey literature search strategy**

The database search strategy was simplified to only include the most central search terms.

To minimise the complexity of the search and thereby limit the number of irrelevant results, the search was run in several steps (four each in Google Search and Google Scholar).

A filter was applied to limit the date range of the search to the year 2000 onwards.

The first 100 results of each single search (N=8) were screened – resulting in 800 screened results overall.

**Search strategy Google:**

1. “mobile clinic” (agriculture OR “domestic work” OR “construction work” OR manufacturing OR mining OR “migrant work”) filetype:pdf
2. “health center” (agriculture OR “domestic work” OR “construction work” OR manufacturing OR mining OR “migrant work”) filetype:pdf
3. “telehealth” (agriculture OR “domestic work” OR “construction work” OR manufacturing OR mining OR “migrant work”) filetype:pdf
4. Low-wage (“mobile clinic” OR “health center” OR telehealth) filetype:pdf

**Search strategy Google Scholar:**

5. “mobile clinic” (agriculture OR “domestic work” OR “construction work” OR manufacturing OR mining OR “migrant work”)
6. “health center” (agriculture OR “domestic work” OR “construction work” OR manufacturing OR mining OR “migrant work”)
7. “telehealth” (agriculture OR “domestic work” OR “construction work” OR manufacturing OR mining OR “migrant work”)
8. Low-wage (“mobile clinic” OR “health center” OR telehealth)

Searches were run between 21^st^ and 23^rd^ of June 2023.
