## Supplement5_StudyOverview for "Healthcare interventions for low-wage migrant workers: A systematic review"

**Supplement 5:** Characteristics of healthcare interventions ordered by type (N=21)

| **First author (Year)** | **Country** | **Type** | **Healthcare provided** | **Time and place of delivery** | **Further services** | **Staff** | **Actors mentioned** | **Funding** | **Patient demographics as reported** | **Work sector** |
| --- | --- | --- | --- | --- | --- | --- | --- | --- | --- | --- |
| Latoo (2021) | Qatar | Hospital in indus-trial area | General hospital services; focus on psychiatric services | 24/7 services at hospital | Mental health training for medical teams of major employers | Nurses, psychiatrists, psychologists, social workers, occupational therapists, dieticians, pharmacist | Government | Government | South Asian, single, male migrants | Industry (craft and manual) |
| Ingram (2015) | USA | Health centre | Primary healthcare | [unclear] | CHWs: health education, facilitating access | Healthcare staff (nursing, internal medicine, paediatrics, obstetrics, medical assistants), clinic staff, CHWs | Federally-qualified health centre | Government | Mexican American farmworkers | Agriculture |
| Garcia (2012) | USA | Health centres | Primary and supplementary healthcare | Across USA | Occupational/ environmental health services, health promotion | Multidisciplinary medical staff | Federally-qualified health centres | Government | Migrant workers (Mexico/Latin America, Asia), mainly young | Mainly agriculture |
| Chen (2010) | China | Health centres | General and occupational health examination | In industrial area, yearly | Workplace inspections, health education (workers and employers), other health promotion | Nurses and physicians trained in occupational health | Government | Government and employers | Mostly (internal) migrant workers | Manufacturing |
| Qian (2007) | China | Factory clinic | First aid, occupational health services | Factory | - | Two doctors | Private factory | Private factory | Unmarried internal female migrants | Manufacturing |
| Heravi (2007) | USA | Week-end clinics | Primary healthcare | Weekend clinics in established medical facilities | Health education, outreach visits, basic health screening | Medical students, volunteer physicians | Local health department, local NGO, university | Student fundraising, university, donations | Migrant farmworkers + families | Agriculture |
| Lukes (2006) | USA | Health centres | General health services, but on dental services | Across the USA; 50% of centres open in evenings | - | Dental healthcare staff (and other healthcare professionals) | Federally-qualified health centre | Government | Migrant farmworkers, mainly Spanish speaking | Agriculture |
| Burgel (2004) | USA | Clinic | Occupational health clinic (screening and basic treatment) | Industrial area, two evenings per month | Health education and other health promotion, meal provided for patients during clinic session | Occupational health nurse+physician, healthcare students, volunteer orthopaedist, clinic staff, interpreter | Local NGOs, university | [unclear] | Migrant workers (Cantonese, female, mostly monolingual), average age: 49 years | Manufacturing |
| Lausch (2003) | USA | Nurse-mana-ged centres | Primary healthcare | Temporary centres (3-6 months during summer) | Health education and promotion | Nurses, bilingual outreach workers, office manager | Federally-qualified health centre | Government | Hispanic migrant farmworkers + families | Agriculture |

**Supplement 5:** Characteristics of healthcare interventions ordered by type (N=21) (continued)

| **First author (Year)** | **Country** | **Type** | **Healthcare provided** | **Time and place of delivery** | **Further services** | **Staff** | **Actors mentioned** | **Funding** | **Patient demographics as reported** | **Work sector** |
| --- | --- | --- | --- | --- | --- | --- | --- | --- | --- | --- |
| Di Gennaro (2021) | Italy | Mobile clinic | Primary healthcare, dental care | At settlements | - | Specialised physician, dentist, nurses, cultural mediator, volunteers, logistician | iNGO | iNGO | Mainly migrant workers, young and male | Agriculture |
| Corwin (2021) | USA | Mobile clinic | Primary healthcare | During COVID-19: evening telephone consultations, optional in-person follow-up | Health and safety training, COVID-19 mitigation; stationary clinic offers comprehensive social services | Bilingual healthcare staff [not specified] | Federally-qualified health centre | Government | Latinx migrant farmworkers, mainly from Mexico/Texas | Agriculture |
| Gruchy (2019) | South Africa | Mobile clinic | HIV, TB, antenatal, maternal, paediatric care | On farms | CHWs leading patient support groups | Nurses | Parallel mobile clinics (iNGO, government) | iNGO, government | Zimbabwean migrant workers | Agriculture |
| Etienne (2016) | Domini-can Republic | Mobile clinic | Primary health-care, dental assessment, paediatrics | In community, 2x/year | Health education, distribution of donated goods | Volunteer nurses, physicians, nursing students, nuns | iNGO, universities in USA and Dom. Republic | iNGO | Mostly Haitian migrant workers + families | Agriculture |
| Hiebert (2015) | Domini-can Republic | Mobile clinic | At least primary healthcare | In community, varying frequency | - | International health professionals | iNGO | iNGO | Mostly Haitian migrant workers + families | Agriculture |
| Luque (2012) | USA | Mobile clinic | Primary healthcare, dental screening, physical therapy | In community, varying frequency | Health education | Health professionals and students, including volunteers | Local NGO, federally-qualified health centre, university | [unclear] | Latino migrant farmworkers, mostly male | Agriculture |
| Brumitt (2011) | USA | Mobile clinic | Primary healthcare, dental examinations | At farms during work | Health education, including stretching | Nurses, other volunteer healthcare professionals, healthcare students | Farm owners, health care staff, university, commu-nity organisations | [unclear] | Latino migrant farmworkers + families | Agriculture |
| Parikh (2010) | Domini-can Republic | Mobile clinic | Primary healthcare, paediatrics | Monthly community visits | Food supplementation | Physicians, CHWs | Local physicians, US university | US university | Mostly Haitian migrant workers + families | Agriculture |
| Crouse (2010) | Domini-can Republic | Mobile clinic | Primary healthcare, paediatrics | Monthly community visits (during the week at daytime) | - | Medical staff, including general physician | Mobile medical team from US | [unclear] | Haitian migrant workers + families | Agriculture |
| Connor (2007; 2010) | USA | Mobile clinic | Primary healthcare, paediatrics, physiotherapy, dental care | Two weeks during peak farming season in evenings at farms/ settlements | Health promotion, childcare during workdays | Health professionals and students, interpreters | Universities, federally-qualified health centre, local NGOs | Universities, local NGOs, stationary farmworker clinic | Migrant farmworkers + families | Agriculture |
| Liem (2022) | China | Tele-health | Mental health app | Flexible | - | E-helpers, clinical supervisors, NGO staff | Academic institutions, local NGOs | University, NGO | Overseas Filipino workers | Mostly domestic workers |
| Price (2013) | USA | Tele-health | App for managing chronic diseases | Flexible | - | [App can facilitate patient provider communication] | University | [research funding] | Hispanic migrant farmworkers, mostly male, young | Agriculture |
